# Drivers of Early Childhood Vaccination Success in Nepal, Senegal, and Zambia: A Multiple Case Study Analysis Using the Consolidated Framework of Implementation Research

**DOI:** 10.1101/2023.04.05.23288208

**Authors:** Cam Escoffery, Emily Ogutu, Zoe Sakas, Kyra A. Hester, Anna Ellis, Katie Rodriguez, Chandni Jaishwal, Chenmua Yang, Sameer Dixit, Anindya Bose, Moussa Sarr, William Kilembe, Robert A. Bednarczyk, Matthew C. Freeman

## Abstract

**Introduction** The fundamental components of a vaccine delivery system are well-documented, but robust evidence is needed on *how* the related processes and implementation strategies - including the facilitators and barriers-contribute to improvements in childhood vaccination coverage. The purpose of this study was to identify critical facilitators and barriers to the implementation of common interventions across three countries that have dramatically increased coverage of early childhood vaccination over the past 20 years, and to qualify common or divergent themes in their success.

**Methods** We conducted 277 key informant interviews and focus group discussions with public health leaders at the regional, district, and local levels and community members in Nepal, Senegal, and Zambia to identify intervention activities and the facilitators and barriers to implementation. We used thematic analysis grounded in the Consolidated Framework for Implementation Research (CFIR) to identify immunization program key facilitators and barriers.

**Results** We found that the common facilitators to program implementation across the countries were the CFIR inner setting constructs of 1) networks and communications, 2) goals and feedback, 3) relative priority, and 4) readiness for implementation; and outer setting constructs of 4) cosmopolitanism and 5) external policies and mandates. The common barriers were incentives and rewards, available resources, access to knowledge and information, and patients needs and resources. Critical to the success of these national immunization programs were prioritization and codification of health as a human right, clear chain of command and shared ownership of immunization, communication of program goals and feedback, offering of incentives at multiple levels, training of staff central to vaccination education, the provision of resources to support the program, key partnerships and guidance on implementation and adoption of vaccination policies.

**Conclusion** Adequate organizational commitment, resources, communication, training, and partnerships were the most critical facilitators for these countries to improve childhood vaccination.

## Introduction

Vaccination averts an estimated 4-5 million deaths annually; children, in particular, have benefited from this protection against communicable diseases [1]. Countries have reported progress in vaccine delivery over the years; however, immunization coverage has varied widely among and within countries [2]. Several countries within Africa and SouthEast Asia regions have outperformed their peers with significant increases in routine immunization coverage since 2000 – including Nepal, Senegal, and Zambia [3–6]. Understanding the factors that contributed to the success of these countries can add to the existing literature and be adopted by other countries to improve their immunization program performance.

To disseminate effective strategies to increase childhood immunization, there needs to be understanding of what works (facilitators) to improve immunization performance among different countries. Identification of those factors can reduce childhood mortality, medical expenditures and increase the future productivity of the country through the longevity of their children. Implementation science is the study of methods to promote the adoption and implementation of evidence-based interventions, strategies, or policies to public health and healthcare settings [7]. This provides us with a framework to help understand the context of implementation, assess and improve public health performance, and support and inform intervention scale-up [8]. The use of implementation science frameworks to examine childhood vaccination programs in countries with high immunization coverage rates provides an opportunity to identify and describe factors that may have supported effective programs and childhood vaccination improvements in low- and middle-income countries (LMICs). The Consolidated Framework for Implementation Research (CFIR) is a widely applied framework that describes the context in which interventions and programs are delivered. CFIR is a meta theory that includes five domains: intervention characteristics, inner setting, outer setting, individual characteristics, and process (Table 1)[9]. It has been widely used since its creation and subsequent publication in 2009 [10], and can explain important factors related to the results of program delivery (e.g., implementation outcomes) [11].

**Table 1.** Inner and Outer Setting Domains of the Consolidated Framework for Implementation Research

| **Consolidated Framework for Implementation Research Constructs** |  |  |
| --- | --- | --- |
| **Construct** |  | **Description** |
| **II. INNER SETTING** |  |  |
| A | Networks & Communications | The nature and quality of webs of social networks and the nature and quality of formal and informal communications within an organization. |
| B | Implementation climate | The absorptive capacity for change, shared receptivity of involved individuals to an intervention, and the extent to which use of that intervention will be rewarded, supported, and expected within their organization. |
|  | B1. Organizational Incentives & Rewards | Extrinsic incentives such as goal-sharing awards, performance reviews, promotions, and raises in salary, and less tangible incentives such as increased stature or respect. |
|  | B2. Goals and Feedback | The degree to which goals are clearly communicated, acted upon, and fed back to staff, and alignment of that feedback with goals. |
|  | B3. Relative Priority | Individuals' shared perception of the importance of the implementation within the organization. |
| C | Readiness for implementation | Tangible and immediate indicators of organizational commitment to its decision to implement an intervention. |
|  | C1. Available Resources | The level of resources dedicated for implementation and on-going operations, including money, training, education, physical space, and time. |
|  | C2. Access to Knowledge & Information | Ease of access to digestible information and knowledge about the intervention and how to incorporate it into work tasks. |

| **I. OUTER SETTING** |  |  |
| --- | --- | --- |
| A | Patient Needs & Resources | The extent to which patient needs, as well as facilitators and barriers to meet those needs, are accurately known and prioritized by the organization. |
| B | Cosmopolitanism | The degree to which an organization is networked with other external organizations. |
| C | External Policy & Incentives | A broad construct that includes external strategies to spread interventions, including policy and regulations (governmental or other central entity), external mandates, recommendations and guidelines, pay-for-performance, collaboratives, and public or benchmark reporting. |

A few studies have applied CFIR to examine children immunization programs internationally among LMICs [12–14]. These studies used CFIR to explain facilitators and barriers to implementation that impacted intervention effectiveness for vaccination initiatives. Facilitators in program delivery included intervention flexibility, self-efficacy of health workers, leadership support and resources (vaccine stocking), and cold chain supervision (structural characteristics). Barriers included acceptability of the vaccine, vaccine costs to mothers, vaccine hesitancy, inadequate cold chain infrastructure and lack of incentives for health workers such as community health workers [12, 15]. A quantitative evaluation study of the Global Polio Eradication Initiative (PEI) in Nigeria using CFIR found that successful contributing factors of community engagement for PEI were the external social environment, and political factors [14]. Application of the CFIR framework may guide the design or selection of more appropriate interventions for a particular health facility or community, identify critical factors at the national and subnational levels of government that lead to better public health delivery. This can ultimately lead to greater acceptability and adoption of vaccination protocols by healthcare providers, which can contribute to increased levels of vaccine uptake [10, 16].

The purpose of this study was to identify common interventions, and the critical facilitators and barriers to implementation using CFIR framework, that contributed to exemplary growth in routine early childhood vaccine coverage in Nepal, Senegal, and Zambia. Findings from this study may be used to develop actionable recommendations for improving immunization programming globally. We examined the CFIR inner setting (i.e., Ministry of Health) and outer setting (i.e, external partners and stakeholders) constructs that were related to vaccine service delivery and demand generation.

### Methods and materials

*Study design*. The Exemplars in Vaccine Delivery research project focused on how Nepal, Senegal, and Zambia succeeded in achieving catalytic growth in childhood routine immunization coverage from 2000-2019. Details about the overall project and methodological approach can be found elsewhere [17, 18]. We used a multi-case study design to explore critical determinants to the implementation of the national immunization programs by applying CFIR, as described in Table 1. Among the five domains of the CFIR framework, we focused on the relevance of the inner and outer settings.

Our specific research questions for this paper were: 1) What are the key intervention strategies employed by the national vaccination programs? and 2) What are the facilitators and barriers related to the implementation of the vaccine programs?

*Procedures*. In consultation with national stakeholders and available data, we selected three provinces within Nepal, Senegal, and Zambia, while considering population density, vaccination coverage, and contextual factors [17]. Qualitative data were collected between August 2019 and April 2021. Data were collected by the Center for Family Health Research in Zambia (CFHRZ), Center for Molecular Dynamics Nepal (CMDN) in Nepal, Institute de Recherche en Santé de Surveillance Epidémiologique et de Formation (IRESSEF) in Senegal, and the Emory University research team. The participants included the Ministry of Health (MoH) staff, key partners, public health officers, local organizations, and community members (Table 2).

**Table 2.** Summary of countries, partner organizations, regions, districts and data Collection Activities[24]

| Country | Nepal |  | Senegal |  | Zambia |  |
| --- | --- | --- | --- | --- | --- | --- |
| In Country Research Partner | Center for Molecular Dynamics, Nepal |  | Institut de Recherche en Santé de Surveillance, de Surveillance Epidémiologique et de Formation (IRESSEF) |  | Center for Family Health Research in Zambia |  |
| Data collection period (MM/YYYY) | 8/2019 – 12/2019 |  | 12/2020 - 4/2021 |  | 10/2019 – 02/2020 |  |
| Regions and Districts |  |  |  |  |  |  |
| Region 1 | Madhes* | Dhanusha, Bara, Mahottari | Ziguinchor | Ziguinchor, Oussouye, Diouloulou | Lusaka | Lusaka, Rufunsa, Chongwe |
| Region 2 | Bagmati | Makwanpur, Dolakha, Kathmandu | Dakar | Rufisque, Mbao, Keur Massar | Central | Chibombo, Chitambo, Serenje |
| Region 3 | Gandaki Pradesh | Kaski, Myagdi, Nawalparasi | Tambacounda | Tambacounda, Koumpentoum, Goudiry | Luapula | Chipili, Nchelenge, Samfya |
| Key Informant Interviews |  |  |  |  |  |  |
| Key: Number of KIIs (Participants) | 79 (79) |  | 62 (63) |  | 66 (85) |  |
| National level government staff | 11 (11) |  | 5 (5) |  | 11 (12) |  |
| Partner organization staff | 8 (8) |  | 4 (4) |  | 11 (15) |  |
| Regional health staff | 5 (5) |  | 7 (7) |  | 6 (8) |  |
| District health staff | 15 (15) |  | 38 (38) |  | 10 (19) |  |
| Health facility staff | 23 (23) |  | 6 (6) |  | 7 (10) |  |
| Community leaders | 15 (15) |  | 2 (2) |  | 10 (10) |  |
| Community health workers ** | 2 (2) |  | - |  | 11 (11) |  |
| Focus Group Discussions |  |  |  |  |  |  |
| Key: Number of FGDs (Participants) | 30 (191) |  | 19 (128) |  | 22 (132) |  |
| Community health workers ** | 9 (60) |  | 10 (65) |  | 10 (60) |  |
| Mothers | 9 (60) |  | 9 (63) |  | 8 (48) |  |
| Fathers | 6 (36) |  | - |  | 1 (6) |  |
| Grandparents | 6 (35) |  | - |  | 3 (18) |  |
| Total (per country) | 109 (270) |  | 81 (191) |  | 88 (217) |  |
| Total (across countries) | - |  | - |  | 278 (678) |  |

*\*Includes volunteer community health workers, female community health volunteers (FCHV),* *vaccinators, bajenu gox, and neighborhood health committee members.*

*Measures.* Topic guides asked participants about facilitators and barriers to implementation, immunization activities and interventions, drivers for high routine immunization coverage, and key partners and their roles. Key informant interview (KII) and focus group discussion (FGD) guides were translated into local languages by research assistants. All interview guides were piloted before use and adjusted iteratively throughout data collection. Data was collected by a different team, other than the authors and the names of the participants were not included. Data collection tools can be found on Open Science Framework [19].

*Data analysis*. Key informant interviews and focus group discussions were audio-recorded. Recordings were transcribed verbatim into the language of the original audio recording, and then translated into English. When audio recordings were in English, the recordings were transcribed directly. Data were password protected, and reviewed by more than one person in the research team, to ensure accuracy and completeness in deidentification. We created a codebook that was iteratively improved based on the emerging themes [20, 21]. Codes were deductive based on CFIR constructs and open-ended questions. Participants were categorized based their role: 1) Ministry at the national level, 2) Ministry at the subnational level, 3) Partners, 4) Local implementers, and 5) Community members. Transcripts were coded and analyzed using MaxQDA2020 software (Berlin, Germany). We used the visual tool in MaxQDA to detect the frequency of codes to select the major CFIR constructs within the inner and outer settings for this analysis. For each theme, we described the valence of the construct to vaccination coverage success: positive influence on implementation, negative, or mixed. At the individual transcript level, valence was determined by interviewees’ accounts related to the specific construct (e.g., available resources, external mandates, etc.). At the case level, we considered whether the interviewees agreed with each other in terms of the constructs’ influence on vaccination. Finally, we assigned an overall valence across the interviewee categories for constructs using methods recommended by others [22].

*Ethics.* The study was considered exempt by the Institutional Review Board committee of Emory University, Atlanta, Georgia, USA (IRB00111474) and approved by ethics committees in each country; Nepal Health Research Council in Kathmandu, Nepal (NHRC; Reg. no. 347/2019); the National Ethical Committee for Health Research (CERNS; Comité National d’Ethique pour la Recherche en Santè) in Dakar, Senegal (00000174); and the University of Zambia Biomedical Research Ethics Committee (Federal Assurance No. FWA00000338, REF. No. 166-2019) and the National Health Research Authority in Zambia. All interviewees gave informed consent to participate in the study.

## Results

We conducted 277 activities in Nepal, Senegal, and Zambia (Table 2). Table 3 outlines key interventions, programs, and policies across all three countries, organized by level of implementation. At the national level, the most commonly implemented interventions across the three countries were the introduction of new vaccines, presence of a national immunization technical advisory board, expansion of health posts, and media campaigns. At the subnational level, all countries reported use of the Reaching Every District (RED) initiative [23], micro-planning, expansion of the vaccine cold chain, community health worker training, monitoring and evaluation meetings, and vaccination campaigns. At the local level, all employed community health workers, outreach efforts for vaccine promotion, media outreach, micro-planning, defaulter tracing, community and religious leadership engagement, and education of grandparents and/or parents using community groups.

**Table 3.** Immunization Programming from Key Informants, Organized by Country and Level of Implementation

| **Level of implementation** | **Policy or intervention** | **Nepal** | **Senegal** | **Zambia** |
| --- | --- | --- | --- | --- |
| **National level programming and policies**<br>*Includes partners and national government* | Introduction of new vaccines | X | X | X |
|  | Strategic reallocation of targeted funds | X |  | X |
|  | Health post expansion | X | X | X |
|  | Codifying health as a human right | X | X |  |
|  | Forums for decision making (e.g., ICC) | X | X | X |
|  | Media engagement | X | X | X |
| **Sub-national programming**<br>*Regional and district level* | RED/REC implementation | X | X | X |
|  | Training CHWs and volunteers | X | X | X |
|  | Meetings for evidence-based decisions | X | X | X |
|  | Data software, health posts |  | X |  |
|  | Data software, district-level | X | X | X |
|  | Cold chain expansion | X | X | X |
| **Community programming**<br>*Health post and village/community level* | Community health worker program | X | X | X |
|  | Community-level health committees | X | X | X |
|  | Outreach services | X | X | X |
|  | Micro-planning in health facilities* | X | X | X |
|  | Defaulter tracing** | X | X | X |
|  | Engagement of community leaders | X | X | X |
|  | Support groups for caregivers | X | X | X |
|  | School outreach | X | X | X |
|  | Promotion through media (e.g., radio) | X | X | X |

\*Micro-planning- integrated set of components prepared to support health system.
\*\*Defaulter tracing- visiting, calling, messaging mothers to remind them about vaccinating their children.

### Facilitators to implementation of vaccination programs

Several CFIR constructs were integral to the implementation of immunization programs across the three countries. Of the 14 CFIR Inner Setting constructs, 3 were found to be facilitators to vaccine delivery: 1) networks and communication, 2) implementation climate, which includes: relative priority, incentives and rewards, goals and feedback, and 3) readiness for implementation, including available resources and access to knowledge. Two CFIR domains - implementation climate and readiness for implementation - were central to the facilitating factors. However, incentives and rewards, available resources, access to knowledge and information, and patients needs and resources constructs were also barriers to implementation of immunization at the local and community level. Table 4 summarizes findings categorized by CFIR constructs and country.

**Table 4.** Salient CFIR Inner and Outer Setting Constructs: Facilitators and Barriers to Implementation of Routine Vaccination Programming

| Construct | Nepal | Senegal | Zambia |
| --- | --- | --- | --- |
| **Inner Setting (Ministry of Health)** |  |  |  |
| **Networks and communication**<br>The nature and quality of webs of social networks and the nature and quality of formal and informal communications within an organization. | + | + | + |
| **Implementation climate** |  |  |  |
| **Relative priority**<br>Individuals' shared perception of the importance of the implementation within the organization. | + | + | + |
| **Incentives and Rewards**<br>Extrinsic incentives such as goal-sharing awards, performance reviews, promotions, and raises in salary, | +/- | +/- | +/- |
| and less tangible incentives such as increased stature or respect. |  |  |  |
| • **Goals and Feedback**<br>The degree to which goals are clearly communicated, acted upon, and fed back to staff, and alignment of that feedback with goals. | + | + | + |
| **Readiness for implementation** |  |  |  |
| • **Available resources**<br>The level of resources dedicated for implementation and on-going operations, including money, training, education, physical space, and time. | +/- | +/- | +/- |
| • **Access to knowledge and information**<br>Ease of access to digestible information and knowledge about the intervention and how to incorporate it into work tasks. | +/- | +/- | +/- |
| **Outer Setting** |  |  |  |
| **Patient Needs and Resources**<br>The extent to which patients' /community members' needs, as well as facilitators and barriers to meet those needs, are accurately known and prioritized by the organization. | +/- | +/- | +/- |
| **Cosmopolitanism** | + | + | + |
| The degree to which an organization is networked with other external organizations (external networks, partners) |  |  |  |
| **External policies and mandates**<br>External strategies to spread interventions, including policy and regulations, external mandates, recommendations and guidelines, pay-for-performance, collaboratives, and public or benchmark reporting. | + | + | + |

+ = Facilitator to implementation. - = Barrier to implementation. +/- = Both facilitator and barrier

### Inner Setting

We found 3 inner setting constructs-1) network and communications, 2) implementation climate (relative priority, incentives and rewards, and goals and feedback), and 3) readiness for implementation (available resources and access to knowledge and information) as salient facilitators to implementation of vaccination programs in Nepal, Senegal, and Zambia (Table 4). A few representative quotes related to these themes can be found below with additional quotes located in Open Science Framework [19].

1. Network and communications

Key informants across all countries viewed communication and coordination between staff at the national and subnational levels as facilitators to successful implementation of childhood vaccinations. Interventions to foster communication and coordination in all three countries included 1) frequent meetings to discuss vaccine data, review and identify improvements, and improve data quality; 2) communication channels between levels of government that support shared ownership of immunization activities; 3) micro-planning at district and community levels to align priorities and tailor strategies; and 4) a clear chain of command that facilitated the flow of information.

> “On the relationship side, it is always the sharing of information, we receive instructions or directives. We try to apply them as much as possible, and in return we make our reports, across the medical region, and we always report the difficulties we find at the peripheral level, on the ground and elsewhere. Whenever we need it, we feedback [from the national level] on everything we had expressed as a point for improvement, recommendation. And from time to time, we still feel that the central level is very close to the peripheral level.” (National Chief Medical Officer, Senegal)

2. Implementation climate

Three sub constructs of implementation climate were facilitators of implementation of the immunization programs. These included relative priority, access to knowledge, and incentives and rewards.

*Relative Priority.* Relative priority is the shared perception of importance of the public health topic and/or program, and it can lead to adoption and/or implementation of public health interventions by program leaders and staff. Public health professionals perceived that there was strong political will and commitment by the government to conduct childhood immunization programming in all three countries. Informants from Senegal and Nepal reported codification of health as a human right in their constitution, and in Zambia, equitable access to quality healthcare as a national priority as included in the current Vision 2030 Zambia strategic document. In Nepal, an informant mentioned that immunization programming was prioritized by the government through implementation of the existing policies that support immunization. In Zambia, there were discussions of having specific budget line for immunization at the national level, showing the level of commitment by the Ministry of Health to improve immunization.

> “I think I may have mentioned this at one point that the government has been procuring the vaccines. In fact, we started own procurement [of vaccines] in 2004. We assumed the total budget for vaccines procurement at that time. A dedicated budget line for vaccine procurement for vaccines was even established within the yellow book. That helped to make sure we had commodities readily available within the facilities. (Former EPI manager, Zambia)

> **“**It is the commitment of government to conduct the program and commitment of policy makers and decision makers, after that commitment of all the health workers working in implementation level. Commitment and involvement of the social organizations and bodies in vaccination program, along with that commitment in the management of vaccines and equipment where it is necessary.” (Former Director General, Nepal)

*Incentives and Rewards*. All three countries reported use of incentives to motivate personnel to improve their performance and increase their vaccination coverage. These included motivating both health workers to improve their staff performance and caregivers to increase their demand for vaccination. In Zambia, supervisors provided trophies to high-performing facilities, districts, and provinces and partners offered non-monetary incentives to communities (e.g., certificates). Health professionals were primarily motivated to vaccinate children to reduce disease among children and ensure healthy communities. Nepal offered incentives at multiple levels: a reward system existed at the district level, and local community health workers were sometimes given a stipend during vaccination campaigns, and they attended training and conferences. Villages, districts and provinces were declared fully immunized and celebrated by the national government through the Full Immunization Declaration (FID) Program in a bottom-up approach, and this motivated local government bodies, service providers, community workers, volunteers and parents and facilitated community buy to immunization. Physicians offered postnatal gifts like mirrors, mosquito nets and other incentives to mothers who got their child vaccinated. In Senegal, recognition was common at different levels, including certificates, upward mobility for high performing facility health workers, and awards and recognition for revenue of health posts.

> “Staff performance is assessed and there’s even an incentive to reward staff if they are performing well at the end of a time. But initially, I think there is every once in a month where we do the Labor Day celebrations where they award the staff that are deserving. So, they awarded five staff that are doing well and they give award just to motivate those staff…” (National Chief Cold Chain Officer, Zambia)

> “We organized for Labor Day; we organized a small gathering. Everyone was there, the service providers, the ASC (community health worker), the bajenu gox, the community actors in general, and there were certificates of recognition that were decided beforehand. We identified the recipients, and these names were put on the certificates. During these gatherings, the doctor gives the certificates along with an envelope of (25,000 francs).” (Sexual and Reproductive Health Coordinator, Senegal)

However, a lack of tangible incentives (money and awards to staff), and inconsistent motivation of community health workers were highlighted as barriers in Nepal and Senegal. Tangible incentives, especially those that were planned in advance (monthly Labor Day celebrations, field allowance or training, for instance) were facilitators.

> “They are not at all motivated, it’s just when there is training that’s the source of motivation or if there is campaign and others but apart from that they have no specific motivation.” (MoH Staff member, Senegal)

> “The health workers are the same who worked previously in health institutions so there are not much differences. However, they don’t get field allowance now that is why motivation level is quite low.” (District Health Officer, Nepal)

*Goals and Feedback*. All three countries had mechanisms for communicating their immunization goals to ensure feedback was shared across all levels. The national level provided instructions, directions, and support for logistics and capacity building. Public health professionals at the subnational level often applied learning from training and logistical support. In all countries, feedback is given from the national to provincial and to the district level at regular intervals (i.e., monthly, annually). In Zambia, Provincial Medical Officers present their program data to the central level on a quarterly basis - including the epidemiology of vaccine preventable diseases - and the national level provides feedback to the provinces. Informants in Nepal stated that feedback is provided to low coverage areas or low performing health facilities, and that their public health staff employ micro-planning and categorization of the health facilities based on that performance. In Senegal, informants indicated that performance was assessed at the start of the month with objectives and targets set for each health post. Feedback on indicators and achievements were provided from the central and provincial levels during monthly meetings.

> “The central level calls the Provincial Medical Officers, on a quarterly basis, sometimes even more often to come and present their data looking at the programs, including EPI, and feedback is given…” (Director of Public Health, Zambia)

> “Recently we have done review at provincial level, which is done yearly. In June each district level data was observed. What is the status level of every district, is everything scheduled? All these were discussed and at the upcoming years what decisions should be made, to make vaccination programs more fruitful what should be done in entire districts. We worked along with family welfare departments and discussion are made. They [family welfare departments] are our responsibilities, and they belong to this group and based on that review is done.” (Immunization supervisor, Nepal)

3. Readiness for implementation

Three sub-constructs of readiness for implementation were perceived as being both facilitators and barriers to the implementation of the immunization programs. These included available resources, access to knowledge, and incentives and rewards.

*Available Resources*. Available resources was a strong theme, and consistent across the countries; this included budgeting for the national immunization programs, cold chain infrastructure, and a vaccination workforce. In Zambia, key informants at the sub-national and local implementation level mentioned availability of staff and volunteers as an enabler for increases in vaccine coverage and sustainability. In addition, cold chain improvements were essential for increases in vaccine coverage and was prioritized by the MOH, who advocated for funding from the Ministry of Finance and from donors to support the infrastructure. In Nepal and Senegal, the government provides the main startup money for the immunization program and there is substantial support by international donor organizations. At the subnational and community levels in Nepal, the local government allocated funds to train and incentivize health workers, maintain health facilities, and hire cold chain technicians. In Senegal, external partners such as GAVI and USAID are mentioned as working at all levels, and some ministry staff noted that they fill in the gap where the district or health development committee cannot fund activities.

> “Basically, the Ministry of Finance decides how much money goes to all the other sectors but when it comes to the EPI, at least we have some specific budget lines and we actually do contribute towards coming up with those plans and budgets but ultimately is the Ministry of Finance that, who determine how much goes to what.” (MoH staff member, Zambia)

> “Now … I cannot say it for sure … but since last two decades, it [vaccine rate] started to show improvement. We can also say after external support started to come like GAVI is our partner … they have played great role, even they help community financially to strengthen system they have played major role in it. They are globally big donor in field of vaccine. They have contributed much in Nepal. …. Talking about partners in Nepal, UN agencies is UNICEF main mandatory, it works mainly for women and children, they also provide technical support to us and WHO are also providing technical support as well.” (National level stakeholder, Nepal)

However, participants at the sub national level reported a lack of available resources which resulted in consistent barriers across the three countries. These constraints included financing for the immunization program, procurement of vaccines, transportation for vaccine provision, human resources, lack of vaccination facilities, and inadequate vaccine supplies.

> “At the district level, there are health posts that are suffering in terms of equipment, and health posts that are suffering from the building. Others are suffering from the resources of transport for the nurses in charge of vaccination.” (MoH staff member, Senegal)

*Access to Knowledge and Information*. Across all countries, training, and quality assurance methods were critical. In Zambia, multiple trainings around the Reaching Every District (RED) were mentioned. RED is a WHO strategy to increase vaccination rates through building capacity to increase vaccine delivery by establishing outreach services, planning and management of resources, monitoring and use of data, and linkage of services with the community [23]. Likewise, Senegal stakeholders discussed that knowledge is passed from the national level to lower levels through training sessions, modules, and manuals. Some district level staff reported organizing and conducting training for community-level workers.

> “The first thing you do is train the districts in RED (Reaching Every District and Every Child). Then they do all the micro plans, everything in the district and we’ll also go and train them. Let me start with, you have to conduct the TOT, the training of trainers. So, we identify the trainers within the districts and provinces. So, eventually we went to train the districts and the districts are going to train the facilities. So, we go and monitor the facilities assuming they are implementing RED and developing all the micro plans and everything. And that is a continuation in other interventions that are coming on board, RED is also a priority. RED is always a training and RED is always a priority.” (Chief Cold Chain Officer, Zambia)

Nevertheless, this construct of access to knowledge was discussed across all countries as a common barrier at the levels closer to the community. There was limited, or infrequent, vaccination training and educational community outreach to all staff, organizations, or partners. Education was sometimes provided to only selected staff, or training information was not disseminated to others in the organization or partners.

> “The training was for one week only. …It was just presentations by a member of staff from the clinic, … The selected group then came here at the clinic to be trained on what to do and say to the people in the community. Yes madam, that is all that they did to train us. (Neighborhood Health Committee member, Zambia)

> “Training is not given much nowadays. The municipality organizes training only once a year.” (Vaccinator, Nepal)

“It may not be very frequent, but we are given training every 6 months.” (Community health worker, Senegal)

### Outer Setting

According to key informants, cosmopolitanism, and external policies and mandates were the two outer setting factors recognized as facilitators for implementation of the vaccine program. Other representative quotes related to these themes are found at Open Science Framework web page [19].

#### Cosmopolitanism

Cosmopolitanism, the extent to which an organization (e.g., the Ministry of Health) is connected to external partnership or networks, was instrumental in implementation. At each country’s national level, there were similar organizations that aided in the implementation of the immunization program. These partners included WHO, UNICEF, and GAVI which offered technical support and financial resources across the three countries. In Nepal, WHO and UNICEF fostered a high trust, constant communication, and collaboration and agreed upon division of labor. At the local level, MoH workers collaborated with religious leaders, schools, NGOs, the media, and community health workers (CHWs) to increase vaccination in all the countries. In Nepal and Zambia, school directors and teachers shared information about immunization with children in school, while Nepal focused on strong media partnerships. In Senegal, community-based organizations and neighborhood delegates supported implementation.

> “In major vaccination, of Polio campaign to support government technically, financially there was WHO and UNICEF. UNICEF majorly supported government in capacity development, logistic management, and in few types of equipment; and after that the global organization such as GAVI, Lions started to support us and inside Nepal Rotary Club, Lions Club started to support us in Polio campaign as a sensitizer and mobilize but in government program there was major support of WHO and UNICEF, where GAVI has constantly supporting us.” (Former Director General, Nepal)

> “Like here at our clinic, letters are written…to different zones to make people aware that there will be a vaccination program, again there are letters written to churches that are close by and also to all those that come to this clinic. So, most women or grandmothers are aware of when to bring children here for vaccination. Posters are also stuck for all to know when the vehicle has past, maybe some are at work, but when they return and look on the trees, walls, and all over the markets, they stick them all over even at the shops…even in schools they announce, school children take the message to parents.” (Grandmothers, Zambia)

#### External Policies and Mandates

External policies and mandates may include political directives, regulations, and external mandates. Many of the immunization efforts are performed within a WHO, GAVI, or UNICEF global policy framework. External agencies gave guidance to country leaders who needed their support and approval to implement policies or adopt new vaccine technologies.

> “Nepal hasn’t become capable of making policies on its own but as it has made an international commitment, WHO guides us to make necessary changes in the policies. Hence, the decisions are made on that basis. Of that, the regulations are formulated, and according to this.” (Ministry of Finance personnel, Nepal)

> “The most important is the ACD plan, which has become the ACE plan (aka - RED/REC). Through this plan of Reaching Every District, it is the activities of the districts and of the health posts that are funded to enable the position to perform.” (EPI and PHC focal point, Senegal)

#### Patient Needs and Resources

Patient needs and resources was reported as a facilitator and barrier to program delivery. This construct identifies whether program implementers are aware of community needs and how they respond to those needs. In all the countries, participants mentioned that they worked together with the healthcare workers to address their needs, and this included the selection of community health workers. These CHWs came from close by communities and understood the needs of communities better. Participants reported that parents in all countries were aware of diseases that could be prevented through childhood vaccination. Participants discussed the changes in infrastructure including better transportation and roads which facilitated access to healthcare. In Senegal, participants reported a mobile strategy where healthcare providers traveled to rural patients to provide vaccination services. Healthcare workers and community actors raised awareness and creating demand for vaccination. Outreach and transportation initiatives were identified as barriers to community needs that the Ministries of Health.

> “By the way, I think there are a lot of people who are aware of different diseases, so they have voluntarily gone to vaccinate their children, they know the interest of vaccination. Certainly, there are people who until now do not believe in vaccination, but there are also us, community actors, who go every day to the population to make the population aware of the interests of vaccination. But also, there are the opinion leaders, who are at the level of neighborhood delegates and the bajenu gox and others who are involved in relation to vaccination.” (Head nurse for Health Post, Senegal)

> “The main reason for the existing good immunization coverage is the awareness among people regarding vaccination, which is given to prevent different diseases. Other reasons are the provision of good quality vaccine, the availability of health workers, accessibility of health services, availability of vaccine, road accessibility. In the past, people had to walk for a day to reach the immunization center, but now due to the transportation facility, people can reach there in an hour or even less.” (MoH staff member, Nepal)

Interviewees identified unmet needs as a barrier to implementation of immunization programming. Community barriers included families facing economic hardship, a lack of knowledge about childhood vaccination, reliance on traditional healers or medicine, vaccine hesitancy, low literacy, and language barriers.

> “And the next thing is about the language. In the Hilly area they have different languages and in the Terai area they have different languages. We make the programs in one language, so it does not match with other areas and people do not understand the language.” (Former Director General, Nepal)

> “It would be nice to explain to mom the benefits of the vaccine her child just took because most moms are illiterate. You have to take the time to talk to them about the vaccine and its benefits because otherwise the mother will be discouraged, she will never come back.” (Mother, Senegal)

## Discussion

Our findings describe *how* Nepal, Senegal, and Zambia achieved high routine immunization coverage through investigating facilitators and barriers to program implementation and how they relate to CFIR constructs. CFIR constructs were critical to understanding the success of these countries in increasing childhood immunization. Although there is no “silver bullet” intervention to enhance early childhood vaccination, our findings point to critical facilitators that exist at national, sub-national, and local levels of vaccine delivery. The application of the CFIR model enabled a comprehensive assessment of implementation context; the use of this framework to assess vaccination programming could be expanded to support adaptation of successful implementation strategies and factors for other countries.

Many facilitators to implementation of the national immunization program were shared by key informants from these exemplar countries. Our findings complement existing literature that supported the use of CFIR to explore factors related to implementation.

Support from international partners-GAVI, WHO, USAID, UNICEF and others, and local community partners-religious leaders, traditional leaders, local media stations, community based organizations and others, in service provision and delivery have led to successful rollout of childhood vaccines in low-income countries [25, 26]. Adoption and implementation of policy initiatives or mandates like Reaching Every District is instrumental in enhancing vaccination coverage. Within these countries, districts are required to submit plans annually to reach every child to receive funding from the ministries for their vaccination activities through assessment of data, identification of problems and creation of a work plan. This strategy is endorsed by WHO and has assisted LMIC in achieving high coverage rates of childhood vaccinations [27]. Although all 3 countries did report that external policies were facilitators, they also reported that access to knowledge, patient needs, and available resources were mixed, and usually negative at the community levels, thus the need for consideration of local context, priorities, and needs alongside external policies.

We identified some common barriers across the countries related to immunization delivery. Some respondents, mostly at regional or local levels, believed that training was missing, not frequent enough, or reached various levels of implementation at national to less local regions. Several studies have demonstrated the impact of training on delivery of immunization programs [15, 28]. Some challenges to vaccination at the community level included language, geography, education or poverty status. In analyzing implementation barriers and strategies in polio eradication initiative in the Democratic Republic of Congo and Ethiopia, Deressa et al. found the common barriers include accessibility issues (population movement, geography); gaps in human resources; supply chain; finance and governance and community hesitancy [29]. The CFIR constructs of available resources, [32, 33].

The findings from this review have various practical implications for how to enhance the implementation of childhood immunization programs. First, prioritization of health or vaccines by ministries or governments, provision of resources for vaccination programs, supplies, and workforce at all levels and facilities are essential to vaccine program implementation [34]. Other qualitative analyses have found insufficient funds and resources and staffing issues as barriers to immunization[35, 36]. Secondly, coordination and engagement of different levels of the country (e.g., ministry, districts/subnational, and local community) and stakeholders was crucial to public education to increase community demand and rollout of the programs [4–6, 37]. Related to this, communication and feedback loops and incentives provided motivation for reaching immunization goals; this has been identified in another study focused on immunization delivery [38]. Third, incentives were helpful in motivating staff and provinces or regions to improve vaccination rates. Finally, partnership engagement at all public health levels and community stakeholder engagement is critical to the success of these programs.

### Strengths and limitations

There were several strengths to this study. CFIR informed the qualitative instruments and identified shared factors across countries to fill a gap in the literature related to optimal implementation of vaccine programs globally. It helped examine factors internal to the ministries and public health programs and also external agents that facilitated successful vaccine delivery. Employing CFIR was a strength because it assists in assessing successful determinants of immunization program implementation and barriers to implementation. The stakeholder groups included in this qualitative study were diverse and included the community level to understand their assessment of critical factors that lead to childhood vaccinations. Often, implementation research focuses on program implementers (e.g., public health leaders, implementation staff) and does not include levels of implementation (regional to local) and community members (intervention recipients). Future research could assess other CFIR domains (i.e., process, characteristics of individuals) and their association with implementation success.

This study has several limitations. The data collection instruments focused on the factors that drove catalytic change and did not elicit policies or interventions that were unsuccessful. We focused on the CFIR constructs and sub-constructs of inner and outer setting post-implementation for the majority of this analysis; recall of activities may be more valid with data collection occurring during vaccine program implementation. The CFIR framework has been updated; future research could explore how other domains of CFIR (e.g., individual, process) and new constructs (e.g., critical incidents, local conditions, etc.) contributed to successful immunization program and outcomes such as sustainment of effective intervention strategies employed by these countries [39]. Using qualitative methods to understand historical events was challenging; interviewees often spoke about current experiences rather than discussing historical factors. However, research assistants probed respondents to reflect on longitudinal changes in the immunization program.

## Conclusion

This multiple case study analysis presents the opportunity to explore implementation science determinants that were critical to the successful implementation of childhood vaccination programming in three countries with high immunization coverage. CFIR’s comprehensive and multifaceted domains help capture the complexities of multilevel interventions to increase childhood vaccination. The use of CFIR helps inform why these countries have high vaccination coverage by describing facilitators and barriers to implementing immunization programming at the national, regional, and local/community level. We identified facilitators such as communication of goals and feedback, offering of incentives at multiple levels, training staff central to vaccination education, providing resources to support the program, and maintaining key partnerships. Public health and governmental staff may bolster direct resources to support the immunization infrastructure, communications, collaborations and incentives to improve vaccination. This study identifies a wide range of facilitators and barriers to implementation of vaccine programs across Nepal, Senegal, and Zambia which contributes to the limited literature on implementation research in global vaccination.

## Data Availability

Additional data can be found https://osf.io/7ys4a/?view_only=739ca7a72f9749118b4aa3d2f7b655d9.

## Acknowledgements

We thank Center for Molecular Dynamics, Nepal (CMDN), the Institut de Recherche en Santé de Surveillance Epidemiologique et de Formation (IRESSEF) in Dakar, Senegal and the Center for Family Health Research in Zambia for their partnership in this study. We gratefully acknowledge the participants who gave their time and insights to help us better understand Nepal’s, Senegal’s, and Zambia’s vaccine delivery system, along with facilitators from the Ministry of Health and Population, Department of Health Services, and the Family Welfare Division of Nepal, Ministry of Health and Social Action in Senegal, and the Ministry of Health in Zambia for supporting this research. In addition, we thank Sarah Chesemore, Anna Rapp, Tove Ryman, and Ethan Wong from the Bill and Melinda Gates Foundation; Kate Buellesbach, Nancy Fullman, Nathaniel Gerthe, Gloria Ikilezi, Caitlyn Mason, David Phillips, and Oliver Rothschild, Jordan-Tate Thomas, and Angela Wang from Gates Ventures for technical support; and the Vaccine Exemplars Research Advisory Group for their insights, specifically Agnes Binagwaho, Laura Craw, Carolina Danovaro, Anuradha Gupta, Heidi Larson, Penelope Masumbu, Kate O’Brien, Helen Rees, Lora Shimp, and Aaron Wallace.

## Data Availability Statement

All relevant data are within the paper and its supporting information files.

## Funding

This work was supported by the Bill & Melinda Gates Foundation, Seattle, WA (OPP1195041) with a planning grant from Gates Ventures, LLC, Kirkland, WA. Bill and Melinda Gates Foundation and Gates Ventures were engaged in the project process, provided context, when necessary, but all research activities and publications are solely those of the authors.

## Competing Interests

The authors declare that they have no known competing financial interests or personal relationships that could have appeared to influence the work reported in this paper.

## Notes

### Competing Interest Statement

The authors have declared no competing interest.

### Author Declarations

The study was considered exempt by the Institutional Review Board committee of Emory University, Atlanta, Georgia, USA (IRB00111474) and approved by ethics committees in each country Nepal Health Research Council in Kathmandu, Nepal (NHRC Reg. no. 347/2019) the National Ethical Committee for Health Research (CERNS Comité National d’Ethique pour la Recherche en Santè) in Dakar, Senegal (00000174) and the University of Zambia Biomedical Research Ethics Committee (Federal Assurance No. FWA00000338, REF. No. 166-2019) and the National Health Research Authority in Zambia.

## References

1. WHO. Immunization 2019 [Available from: https://www.who.int/news-room/facts-in-pictures/detail/immunization.

2. WHO. Immunization Agenda 2030 A global strategy to leave no one behind. 2020.

3. WHO. WHO-UNICEF estimates of DTP3 coverage 2020 [updated 15-Jul-2020; cited 2021 23 November]. Available from: https://apps.who.int/immunization_monitoring/globalsummary/timeseries/tswucoveragedtp3.html.

4. Micek K, Hester KA, Chanda C, Darwar R, Dounebaine B, Ellis AS, et al. Critical success factors for routine immunization performance: A case study of Zambia 2000 to 2018. Vaccine X. 2022;11:100166.

5. Hester KA, Sakas Z, Ellis AS, Bose AS, Darwar R, Gautam J, et al. Critical success factors for high routine immunization performance: A case study of Nepal. Vaccine: X. 2022;12:100214-.

6. Sakas Z, Hester KA, Rodriguez K, Diatta SA, Ellis AS, Gueye DM, et al. Critical success factors for high routine immunization performance: A case study of Senegal. medRxiv. 2022:2022.01.25.22269847.

7. Eccles MP, Mittman BS. Welcome to Implementation Science. Implementation Science. 2006;1(1):1.

8. Peters DH, Adam T, Alonge O, Agyepong IA, Tran N. Implementation research: what it is and how to do it. BMJ : British Medical Journal. 2013;347:f6753.

9. Damschroder LJ, Aron DC, Keith RE, Kirsh SR, Alexander JA, Lowery JC. Fostering implementation of health services research findings into practice: a consolidated framework for advancing implementation science. Implementation Science. 2009;4(1):50.

10. Means AR, Kemp CG, Gwayi-Chore M-C, Gimbel S, Soi C, Sherr K, et al. Evaluating and optimizing the consolidated framework for implementation research (CFIR) for use in low-and middle-income countries: a systematic review. Implementation Science. 2020;15(1):1–19.

11. Nilsen P. Making sense of implementation theories, models, and frameworks. Implementation Science 30: Springer; 2020. p. 53-79.

12. Adamu AA, Uthman OA, Gadanya MA, Wiysonge CS. Using the consolidated framework for implementation research (CFIR) to assess the implementation context of a quality improvement program to reduce missed opportunities for vaccination in Kano, Nigeria: a mixed methods study. Hum Vaccin Immunother. 2020;16(2):465-75.

13. Boisson A, Thompson P, Fried B, Shea CM, Ngimbi P, Lumande F, et al. Determinants of the uptake of childhood immunization in Kinshasa Province, the DRC: ordered logit regression analyses to assess timely infant vaccines administered at birth and six-weeks of age. 2022.

14. Akinyemi OO, Adebayo A, Bassey C, Nwaiwu C, Kalbarczyk A, Fatiregun AA, et al. Assessing community engagement in Nigeria polio eradication initiative: application of the Consolidated Framework for Implementation Research. BMJ open. 2021;11(8):e048694.

15. Boisson A, Morgan CE, Fried B, Shea CM, Yotebieng M, Ngimbi P, et al. Barriers and facilitators to timely birth-dose vaccines in Kinshasa Province, the DRC: a qualitative study. Journal of Global Health Reports. 2022;6:e2022028.

16. Selove R, Foster M, Mack R, Sanderson M, Hull PC. Using an Implementation Research Framework to Identify Potential Facilitators and Barriers of an Intervention to Increase HPV Vaccine Uptake. J Public Health Manag Pract. 2017;23(3):e1–e9.

17. Bednarczyk RA, Hester KA, Dixit SM, Ellis AS, Escoffery C, Kilembe W, et al. Protocol: Identification and evaluation of critical factors in achieving high and sustained childhood immunization coverage in selected low- and lower-middle income countries. medRxiv. 2021:2021.12.01.21267018.

18. Health EiG. Making Better Decisions in Global Health: Understand Positive Outliers to Inform Policy and Practice. 2023 [Available from: https://www.exemplars.health/.

19. OSF. Exemplars in Vaccine Delivery2022.

20. Hennink MM. Focus group discussions: Oxford University Press; 2013.

21. Miles MB, Huberman AM, Saldana J. Qualitative Data Analysis: A Methods Sourcebook: SAGE Publications; 2019.

22. Damschroder LJ, Lowery JC. Evaluation of a large-scale weight management program using the consolidated framework for implementation research (CFIR). Implementation Science. 2013;8(1):51.

23. WHO. Essential Programme on Immunization: Reaching Every District (RED) 2023 [Available from: https://www.who.int/teams/immunization-vaccines-and-biologicals/essential-programme-on-immunization/implementation/reaching-every-district-(red).

24. Sakas Z, Hester KA, Ellis AS, Ogutu EA, Rodriguez K, Bednarczyk RA, et al. Critical success factors for high routine immunization performance: A multiple case study analysis of Nepal, Senegal, and Zambia. medRxiv. 2022:2022.11.08.22282076.

25. Gatera M, Bhatt S, Ngabo F, Utamuliza M, Sibomana H, Karema C, et al. Successive introduction of four new vaccines in Rwanda: High coverage and rapid scale up of Rwanda’s expanded immunization program from 2009 to 2013. Vaccine. 2016;34(29):3420–6.

26. Ahonkhai AA, Odusanya OO, Meurice FP, Pierce LJ, Durojaiye TO, Alufohai EF, et al. Lessons for strengthening childhood immunization in low-and middle-income countries from a successful public-private partnership in rural Nigeria. International Health. 2022;14(6):632–8.

27. WHO. Microplanning for immunization service delivery using the Reaching Every District (RED) strategy. World Health Organization; 2009.

28. Mpabalwani E, Menon J, Phiri G, Malambo A, Mbozi E, Kalesha P, et al. Assessing the delivery and effectiveness of a new immunisation training initiative at district level in Zambia. Medical Journal of Zambia. 2011;38(1):8–12.

29. Deressa W, Kayembe P, Neel AH, Mafuta E, Seme A, Alonge O. Lessons learned from the polio eradication initiative in the Democratic Republic of Congo and Ethiopia: analysis of implementation barriers and strategies. BMC Public Health. 2020;20(4):1807.

30. Escoffery C, Riehman K, Watson L, Priess AS, Borne MF, Halpin SN, et al. Facilitators and Barriers to the Implementation of the HPV VACs (Vaccinate Adolescents Against Cancers) Program: A Consolidated Framework for Implementation Research Analysis. Prev Chronic Dis. 2019;16:E85.

31. Soi C, Gimbel S, Chilundo B, Muchanga V, Matsinhe L, Sherr K. Human papillomavirus vaccine delivery in Mozambique: identification of implementation performance drivers using the Consolidated Framework for Implementation Research (CFIR). Implementation Science. 2018;13(1):151.

32. Lane S, MacDonald NE, Marti M, Dumolard L. Vaccine hesitancy around the globe: Analysis of three years of WHO/UNICEF Joint Reporting Form data-2015-2017. Vaccine. 2018;36(26):3861–7.

33. Cooper S, Betsch C, Sambala EZ, McHiza N, Wiysonge CS. Vaccine hesitancy – a potential threat to the achievements of vaccination programmes in Africa. null. 2018;14(10):2355–7.

34. LaFond A, Kanagat N, Steinglass R, Fields R, Sequeira J, Mookherji S. Drivers of routine immunization coverage improvement in Africa: findings from district-level case studies. Health Policy Plan. 2015;30(3):298–308.

35. Ezezika O, Mengistu M, Opoku E, Farheen A, Chauhan A, Barrett K. What are the barriers and facilitators to polio vaccination and eradication programs? A systematic review. PLOS Global Public Health. 2022;2(11):e0001283.

36. Nkwenkeu SF, Jalloh MF, Walldorf JA, Zoma RL, Tarbangdo F, Fall S, et al. Health workers’ perceptions and challenges in implementing meningococcal serogroup a conjugate vaccine in the routine childhood immunization schedule in Burkina Faso. BMC public health [Internet]. 2020 2020/02//; 20(1):[254 p.]. Available from: http://europepmc.org/abstract/MED/32075630 https://doi.org/10.1186/s12889-020-8347-z https://europepmc.org/articles/PMC7031928 https://europepmc.org/articles/PMC7031928?pdf=render.

37. Sakas Z, Hester K, Ellis A, Ogutu EA, Rodriguez K, Bednarczyk R, et al. Critical success factors for high routine immunization performance: A multiple case study analysis of Nepal, Senegal, and Zambia. medRxiv; 2022.

38. Guan TH, Htut HN, Davison CM, Sebastian S, Bartels SA, Aung SM, et al. Implementation of a neonatal hepatitis B immunization program in rural Karenni State, Myanmar: A mixed-methods study. PLoS One. 2021;16(12):e0261470.

39. Damschroder LJ, Reardon CM, Opra Widerquist MA, Lowery J. Conceptualizing outcomes for use with the Consolidated Framework for Implementation Research (CFIR): the CFIR Outcomes Addendum. Implementation Science. 2022;17(1):7.

